# Physical complaints and their relationships to esports activities among Japanese esports players: A cross-sectional study

**DOI:** 10.1101/2023.11.14.23298495

**Authors:** Takafumi Monma, Takashi Matsui, Shoya Koyama, Hiromasa Ueno, Junichi Kagesawa, Chisato Oba, Kentaro Nakamura, Hideki Takagi, Fumi Takeda

**Affiliations:** Institute of Health and Sport Sciences, University of Tsukuba, Ibaraki, Japan; R&D Center for Sport Innovation, University of Tsukuba, Ibaraki, Japan; R&D Center for Lifestyle Innovation, University of Tsukuba, Ibaraki, Japan; REJECT Inc., Tokyo, Japan; NTTe-Sports Inc., Tokyo, Japan; Food Microbiology and Function Research Laboratories, R&D Division, Meiji Co., Ltd., Tokyo, Japan

**Keywords:** electronic sports, video game, somatic symptom, musculoskeletal disease

## Abstract

In the evolving landscape of electronic sports (esports), where economic and social expectations are soaring, a critical concern has emerged in physical complaints among esports players. However, empirical insights into these complaints’ prevalence and influencing factors are scarce. This study aimed to clarify the prevalence of physical complaints and their association with esports activities among Japanese esports players. A web-based cross-sectional survey encompassing 175 esports players from both professional and amateur teams in Japan was conducted. The analysis focused on 79 male participants (average age: 21.6 ± 5.6 years) with complete responses. The survey items included the esports factors (the device mainly used, the duration of esports titles played primarily, hours of esports activities per day on weekdays and holidays, and the distance between the screen and the face during esports activities) and physical complaints (headache, neck pain, stiff or sore shoulders, wrist pain, finger pain, lower back pain, and eye fatigue). A total of 49.4% reported stiff or sore shoulders, 48.1% faced eye fatigue, and 30.4% had headaches. Professionals exhibited a significantly higher likelihood of neck, wrist, and lower back pain and eye fatigue than amateurs. Age-adjusted logistic regression analysis uncovered that using mainly mobile devices and being closer to the screen and face during esports activities were significantly associated with neck pain, stiff or sore shoulders, lower back pain, and eye fatigue. These results suggest that poor posture caused by using mobile devices and being closer to the screen during esports activities was related to various physical complaints.

## Introduction

Electronic sports (esports) is “an organized and competitive approach to playing computer games”^1^ through various platforms (such as consoles, computers, mobile devices, and virtual reality). The global esports audience and active esports players increase every year.^2^ Esports was accepted as medal events in the 19th Asian Games held in 2023,^3^ and the International Olympic Committee officially launched its Olympic Esports Series in 2023.^4^ In Japan, esports events have been the programs of the National Sports Festival since 2019,^5^ and expectations of esports’ economic benefits and social significance are growing.^6^

However, the growing interest in esports has resulted in some considerable downsides. Physical health is a major concern. Excessive video game playing is associated with physical complaints.^7,8^ In addition, previous studies have investigated the prevalence of physical complaints among competitive esports players. DiFrancisco-Donoghue et al. reported that 56% of collegiate esports players in the United States and Canada suffered from eye fatigue, 42% had back or neck pain, 36% had wrist pain, and 32% had hand pain.^9^ Lindberg et al. investigated musculoskeletal pain in esports players trained with a coach in Denmark. They reported that the primary pain sites included the back (31.3%), neck (11.3%), and shoulders (11.3%).^10^ Another study conducted in China on professional mobile esports players reported that the neck (40%) was the most prevalent site of musculoskeletal complaints, followed by the fingers (38%), the head (32%), and the lower back (20%).^11^ Furthermore, a study of competitive video gaming in Saudi Arabia showed that the most reported sites of injuries included the lower back (63.8%), neck (50%), hand/wrist (44.8%), and shoulder (35.3%).^12^ The proportion of esports athletes with physical complaints in Japan is thought to be similar to that of these previous findings, but no empirical study has been conducted.

Furthermore, differences in the proportion of those with physical complaints by attributes such as age, body size, and esports level (professional or amateur) have not been examined among esports players. In general, the proportion of people with physical complaints generally varies by attributes such as age and body size.^13,14^ Furthermore, similar to traditional sports,^15^ there may be differences in musculoskeletal conditions between professionals, who have their periodized training schedules, access to team resources, and season-based play, and amateurs.^16^

The issue of physical complaints in esports players is the same as in the context of occupational health; esports practice requires long periods of computer and console use while sitting in poor postures, repetitive or forceful movements, and prolonged screen exposure.^17–22^ However, few empirical studies have examined the association between esports activities and physical complaints among esports players. A cross-sectional study reported that esports players with musculoskeletal pain participated in less esports training than those without pain.^10^ A study in Saudi Arabia showed that the duration of playing video games and the days played per month were inversely associated with musculoskeletal disorders, with gamers who played for 1–5 years showing a higher rate of musculoskeletal disorders than those who played for more than 10 years.^12^ Another study reported that career duration was not significantly associated with ease of eyestrain or leg numbness.^11^ Considering the previous studies, the duration of esports play and the amount of time played per day may have little relationship with physical complaints. However, other factors of esports activities, such as the device used for esports and posture during play, have not been empirically examined in esports players, and these may be related to physical complaints in light of the points made above.

Therefore, this study aimed to clarify the prevalence of physical complaints and their association with esports activities among Japanese esports players.

## Methods

### Procedure and participants

In this study, representatives from one professional team and five amateur teams were asked to survey all their team members. In total, 175 esports players were surveyed. An anonymous self-administered web-based questionnaire survey was conducted from April to May 2021. The URL of the questionnaire was provided to the representative on April 15 for the professional team and April 14 for the amateur teams. Next, the esports players responded to an anonymous self-administered web-based questionnaire by May 12 for the professional team and May 23 for the amateur teams. One hundred responses were collected (recovery rate: 57.1%). Of these, no women were on the professional team, and only 19 were on the amateur teams. Therefore, women were excluded from the analysis. Furthermore, two men were excluded from the analysis due to incomplete responses. Finally, 79 men with complete responses were included in the analysis.

This study was approved by the Ethics Committee of XXXX [name of the institution is hidden due to double blind review policy], in compliance with the Declaration of Helsinki (Reference No: Tai 021-112). All respondents provided informed consent to participate in the study.

### Questionnaires

The questionnaire included questions on attributes, esports activities, and physical complaints. The attributes included age, height, weight, and team affiliation. Body mass index (BMI) was calculated from height and weight; under 18.5 was considered “underweight,” 18.5 to under 25 as “normal weight,” and 25 or above as “overweight or obese.” Furthermore, the esports level was categorized as professional or amateur based on the teams to which they belonged.

Esports activities included the esports title played primarily, duration of esports titles played primarily, hours of esports activities per day on weekdays and holidays, and the distance between the screen and the face during esports activities. The device mainly used was categorized as “PC or console” and “mobile,” based on the response of the esports title played primarily.

Regarding physical complaints, the following seven items were established that had been prevalent among esports players in previous studies:^9–12^ headache, neck pain, stiff or sore shoulders, wrist pain, finger pain, lower back pain, and eye fatigue. Respondents were asked about the presence or absence of each item.

### Statistical analysis

The esports activities and the presence of each physical complaint were calculated and compared by age (divided by the median), BMI categories, and esports level (professional or amateur) using Fisher’s exact tests, Mann–Whitney U tests, and Kruskal–Wallis tests. After confirming the correlation between each factor of esports activities using Spearman’s rank correlation analysis, the association between esports activities and each physical complaint was investigated using logistic regression analysis. Two logistic regression analysis models were applied: Model 1 was adjusted for age, and Model 2 was adjusted for age and esports level (professional and amateur). The logistic regression analysis used physical complaints reported by 10% or more of the respondents. The distance between the screen and the face during esports activities was converted to 10 cm to avoid an extremely small range of odds ratios. SPSS Statistics 29 for Windows was used for all statistical analyses, and the statistical significance level was set at 5%.

## Results

Table 1 presents the characteristics of the respondents. The mean age was 21.6 ± 5.6 years. Based on BMI, 27 respondents (34.2%) were underweight, 43 (54.4%) were normal weight, and 9 (11.4%) were overweight or obese. Of the respondents, 39 were professionals (49.4%), and 40 were amateurs (50.6%).

**TABLE 1.** RESPONDENT CHARACTERISTICS.

| | | Mean ±SD<br>or n (%) |
| --- | --- | --- |
| Attributes |  |  |
| Age | | 21.6±5.6 |
| Body mass index | Underweight | 27(34.2) |
|  | Normal weight | 43(54.4) |
|  | Overweight or obese | 9(11.4) |
| Esports level | Professional | 39(49.4) |
|  | Amateur | 40(50.6) |
| Esports activities |  |  |
| Esports title played primarily | VALORANT | 5(6.3) |
|  | Rainbow Six Siege | 7(8.9) |
|  | PUBG | 5(6.3) |
|  | APEX Legends | 8(10.1) |
|  | Call of Duty | 3(3.8) |
|  | Call of Duty mobile | 5(6.3) |
|  | PUBG mobile | 7(8.9) |
|  | PUBG mobile (Academy) | 3(3.8) |
|  | League of Legends Wild Rift | 4(5.1) |
|  | Fortnite | 32(40.5) |
| Duration of esports titles played primarily (years) | | 2.4±1.5 |
| Hours of esports activities per day | Weekdays | 4.2±2.9 |
| | Holidays | 4.1±3.3 |
| Distance between the screen and the face during esports activities (cm) | | 44.7±26.7 |
| Physical complaints (Presence) |  |  |
|  | Headache | 24(30.4) |
|  | Neck pain | 23(29.1) |
|  | Stiff or sore shoulders | 39(49.4) |
|  | Wrist pain | 5(6.3) |
|  | Finger pain | 3(3.8) |
|  | Lower back pain | 23(29.1) |
|  | Eye fatigue | 38(48.1) |

Regarding the device, 60 respondents (75.9%) mainly used PCs or consoles, and 19 (24.1%) used mobiles. The mean duration of esports titles played primarily was 2.4 ± 1.5 years, and the mean number of hours of esports activities per day was 4.2 ± 2.9 hours on weekdays and 4.1 ± 3.3 hours on holidays. The mean distance between the screen and the face during esports activities was 44.7 ± 26.7 cm.

Among physical complaints, stiff or sore shoulders were the most common complaints, with 39 respondents (49.4%), followed by eye fatigue with 38 respondents (48.1%), headache with 24 respondents (30.4%), and neck pain and lower back pain with 23 respondents (29.1%) each.

Table 2 shows the esports activities, and the proportion of each physical complaint was assessed according to age, BMI categories, and esports level (professional or amateur). Professionals were more likely than amateurs to use mainly mobile devices (p < 0.001). Furthermore, professionals spent significantly more time on esports activities on weekdays (p < 0.001) and had a significantly shorter distance between the screen and face during esports activities than amateurs (p < 0.001). Regarding physical complaints, professionals were significantly more likely to experience neck pain (p < 0.05), wrist pain (p < 0.05), lower back pain (p < 0.05), and eye fatigue (p < 0.01) than amateurs. There was no difference in esports activities or physical complaints according to age or BMI.

**TABLE 2.** ESPORTS ACTIVITIES AND PHYSICAL COMPLAINTS BY ATTRIBUTES.

|  |  | Age |  |  | Body mass index |  |  |  | Esports level |  |  |
| --- | --- | --- | --- | --- | --- | --- | --- | --- | --- | --- | --- |
|  |  | <20 years | ≥20 years |  | Underweig<br>ht | Normal<br>weight | Overweight<br>or obese |  | Professional | Amateur |  |
|  |  | n (%) or<br>Mean ±SD | n (%) or<br>Mean ±SD | p | n (%) or<br>Mean ±SD | n (%) or<br>Mean ±SD | n (%) or<br>Mean ±SD | p | n (%) or<br>Mean ±SD | n (%) or<br>Mean ±SD | p |
| Esports activities |  |  |  |  |  |  |  |  |  |  |  |
| Device of the esports<br>title mainly played | PC or<br>console | 26(70.3) | 34(81.0) | 0.301<sup>a</sup> | 18(66.7) | 34(79.1) | 8(88.9) | 0.381<sup>a</sup> | 20(51.3) | 40(100.0) | <0.001<sup>a</sup> |
|  | Mobile | 11(29.7) | 8(19.0) |  | 9(33.3) | 9(20.9) | 1(11.1) |  | 19(48.7) | 0(0.0) |  |
| Duration of esports titles<br>played primarily (years) |  | 2.2±1.1 | 2.6±1.8 | 0.425<sup>b</sup> | 2.5±1.9 | 2.5±1.2 | 2.1±1.9 | 0.447<sup>c</sup> | 2.5±1.9 | 2.3±1.1 | 0.906<sup>b</sup> |
| Hours of esports<br>activities per day | Weekdays | 3.7±2.5 | 4.7±3.2 | 0.162<sup>b</sup> | 4.4±2.1 | 4.1±3.4 | 4.2±3.0 | 0.693<sup>c</sup> | 5.9±2.5 | 2.6±2.3 | <0.001<sup>b</sup> |
|  | Holidays | 4.0±3.5 | 4.1±3.2 | 0.835<sup>b</sup> | 4.8±3.5 | 3.7±3.2 | 3.7±3.0 | 0.433<sup>c</sup> | 4.4±2.9 | 3.8±3.7 | 0.334<sup>b</sup> |
| Distance between the<br>screen and the face<br>during esports activities<br>(cm) |  | 45.2±29.3 | 44.3±24.6 | 0.906<sup>b</sup> | 38.5±29.8 | 48.2±24.9 | 46.7±24.6 | 0.093<sup>c</sup> | 26.8±14.1 | 62.1±24.6 | <0.001<sup>b</sup> |
| Physical complaints |  |  |  |  |  |  |  |  |  |  |  |
| Headache | Presence | 10(27.0) | 14(33.3) | 0.628<sup>a</sup> | 7(25.9) | 15(34.9) | 2(22.2) | 0.723<sup>a</sup> | 12(30.8) | 12(30.0) | 1.000<sup>a</sup> |
| Neck pain | Presence | 8(21.6) | 15(35.7) | 0.217<sup>a</sup> | 10(37.0) | 11(25.6) | 2(22.2) | 0.633<sup>a</sup> | 16(41.0) | 7(17.5) | 0.027<sup>a</sup> |
| Stiff or sore shoulders | Presence | 15(40.5) | 24(57.1) | 0.178<sup>a</sup> | 13(48.1) | 23(53.5) | 3(33.3) | 0.621<sup>a</sup> | 23(59.0) | 16(40.0) | 0.117<sup>a</sup> |
| Wrist pain | Presence | 2(5.4) | 3(7.1) | 1.000<sup>a</sup> | 10(37.0) | 12(27.9) | 1(11.1) | 0.408<sup>a</sup> | 5(12.8) | 0(0.0) | 0.026<sup>a</sup> |
| Finger pain | Presence | 1(2.7) | 2(4.8) | 1.000<sup>a</sup> | 3(11.1) | 2(4.7) | 0(0.0) | 0.692<sup>a</sup> | 2(5.1) | 1(2.5) | 0.615<sup>a</sup> |
| Low back pain | Presence | 14(37.8) | 9(21.4) | 0.139<sup>a</sup> | 2(7.4) | 1(2.3) | 0(0.0) | 0.355<sup>a</sup> | 16(41.0) | 7(17.5) | 0.027<sup>a</sup> |
| Eye fatigue | Presence | 15(40.5) | 23(54.8) | 0.261<sup>a</sup> | 14(51.9) | 20(46.5) | 4(44.4) | 0.949<sup>a</sup> | 25(64.1) | 13(32.5) | 0.007<sup>a</sup> |

*Note.* <sup>a</sup> Fisher’s exact test.
<sup>b</sup> Mann–Whitney U test.
<sup>c</sup> Kruskal–Wallis test.

Table 3 presents the correlations between each factor of esports activities. The device mainly used had a significant correlation with the hours of esports activities per day on weekdays (ρ = 0.266, p < 0.05) and the distance between the screen and face during esports activities (ρ = -0.598, p < 0.001). Hours of esports activities per day on weekdays also had a significant correlation with hours of esports activities per day on holidays (ρ = 0.524, p < 0.001) and the distance between the screen and the face during esports activities (ρ = -0.499, p < 0.001).

**TABLE 3.** CORRELATION BETWEEN ESPORTS ACTIVITIES.

|  | (1) | (2) | (3) | (4) |
| --- | --- | --- | --- | --- |
| (1) Device mainly used |  |  |  |  |
| (2) Duration of esports titles played primarily | -0.208 |  |  |  |
| (3) Hours of esports activities per day on weekdays | 0.266* | 0.011 |  |  |
| (4) Hours of esports activities per day on holidays | 0.100 | -0.068 | 0.524** |  |
| (5) Distance between the screen and the face during esports activities | -0.598** | 0.073 | -0.499** | -0.160 |

*Note.* Spearman's rank correlation coefficients.
\* $p < 0.05$ , \*\* $p < 0.001$ .

Table 4 shows the relationships between esports activities and each physical complaint reported by 10% or more of the respondents. In Model 1 adjusted for age, the device mainly used was significantly related to neck pain (OR: 5.62, 95% CI: 1.72-18.33, p < 0.01), stiff or sore shoulders (OR: 7.63, 95% CI: 2.16–27.03, p < 0.01), low back pain (OR: 4.71, 95% CI: 1.52–14.61, p < 0.01), and eye fatigue (OR: 4.45, 95% CI: 1.37–14.41, p < 0.05). The distance between the screen and the face during esports activities also had a significant association with neck pain (OR: 0.61, 95% CI: 0.44–0.85, p < 0.01), stiff or sore shoulders (OR: 0.69, 95% CI: 0.54–0.87, p < 0.01), low back pain (OR: 0.75, 95% CI: 0.59–0.96, p < 0.05), and eye fatigue (OR: 0.71, 95% CI: 0.56–0.89, p < 0.01).

**TABLE 4.** RELATIONSHIPS BETWEEN ESPORTS ACTIVITIES AND PHYSICAL ACTIVITIES.

|  |  | Headache |  |  | Neck pain |  |  | Stiff or sore shoulders |  |  | Lower back pain |  |  | Eye fatigue |  |  |
| --- | --- | --- | --- | --- | --- | --- | --- | --- | --- | --- | --- | --- | --- | --- | --- | --- |
|  |  | OR | 95%CI | p | OR | 95%CI | p | OR | 95%CI | p | OR | 95%CI | p | OR | 95%CI | p |
| Model 1<sup>a</sup> |  |  |  |  |  |  |  |  |  |  |  |  |  |  |  |  |
| Device mainly used (0 = PC or console; 1 = mobile) |  | 0.81 | 0.25-2.65 | 0.723 | 5.62 | 1.72-18.33 | 0.004 | 7.63 | 2.16-27.03 | 0.002 | 4.71 | 1.52-14.61 | 0.007 | 4.45 | 1.37-14.41 | 0.013 |
| Duration of esports titles played primarily |  | 1.07 | 0.79-1.46 | 0.648 | 1.17 | 0.85-1.60 | 0.337 | 1.03 | 0.77-1.37 | 0.865 | 1.12 | 0.81-1.54 | 0.497 | 0.83 | 0.61-1.12 | 0.219 |
| Hours of esports activities per day | Weekdays | 0.98 | 0.83-1.16 | 0.813 | 1.09 | 0.91-1.29 | 0.343 | 0.98 | 0.84-1.15 | 0.804 | 1.20 | 1.00-1.44 | 0.056 | 1.02 | 0.87-1.20 | 0.777 |
|  | Holidays | 0.96 | 0.83-1.12 | 0.631 | 0.94 | 0.81-1.10 | 0.464 | 0.94 | 0.82-1.08 | 0.410 | 0.93 | 0.79-1.08 | 0.339 | 0.92 | 0.80-1.06 | 0.245 |
| Distance between the screen and the face during esports activities |  | 0.91 | 0.75-1.12 | 0.372 | 0.61 | 0.44-0.85 | 0.003 | 0.69 | 0.54-0.87 | 0.002 | 0.75 | 0.59-0.96 | 0.024 | 0.71 | 0.56-0.89 | 0.003 |
| Model 2<sup>b</sup> |  |  |  |  |  |  |  |  |  |  |  |  |  |  |  |  |
| Device mainly used (0 = PC or console; 1 = mobile) |  | 0.69 | 0.17-2.80 | 0.606 | 3.47 | 0.89-13.53 | 0.073 | 7.42 | 1.73-31.78 | 0.007 | 3.42 | 0.84-13.90 | 0.085 | 2.44 | 0.62-9.59 | 0.203 |
| Duration of esports titles played primarily |  | 1.07 | 0.79-1.46 | 0.656 | 1.13 | 0.82-1.56 | 0.456 | 1.01 | 0.75-1.36 | 0.965 | 1.10 | 0.80-1.51 | 0.577 | 0.79 | 0.58-1.08 | 0.136 |
| Hours of esports activities per day | Weekdays | 0.96 | 0.78-1.18 | 0.700 | 0.93 | 0.75-1.16 | 0.546 | 0.85 | 0.69-1.04 | 0.110 | 1.11 | 0.89-1.38 | 0.349 | 0.84 | 0.68-1.04 | 0.106 |
|  | Holidays | 0.96 | 0.83-1.12 | 0.624 | 0.92 | 0.78-1.09 | 0.348 | 0.93 | 0.80-1.08 | 0.338 | 0.91 | 0.76-1.07 | 0.257 | 0.90 | 0.77-1.05 | 0.167 |
| Distance between the screen and the face during esports activities |  | 0.87 | 0.66-1.13 | 0.292 | 0.63 | 0.43-0.94 | 0.024 | 0.65 | 0.48-0.88 | 0.006 | 0.81 | 0.59-1.11 | 0.190 | 0.76 | 0.57-1.01 | 0.056 |

*Note.* <sup>a</sup> Logistic regression analysis adjusted for age.
<sup>b</sup> Logistic regression analysis adjusted for age and esports level.
OR, odds ratio, CI, confidence interval.

In Model 2 adjusted for age and esports level, the device mainly used was still significantly associated with stiff or sore shoulders (OR: 7.42, 95% CI: 1.73–31.78, p < 0.01). Furthermore, the distance between the screen and the face during esports activities had a significant association with neck pain (OR: 0.63, 95% CI: 0.43–0.94, p < 0.05) and stiff or sore shoulders (OR: 0.65, 95% CI: 0.48–0.88, p < 0.01).

## Discussion

This study aimed to clarify the prevalence of physical complaints and their association with esports activities among Japanese esports players. The results showed that stiff or sore shoulders were the most common, followed by eye fatigue, headache, neck pain, and lower back pain. These proportions were similar to previous studies.^9,11^ In contrast, wrist and finger pain was reported as considerably less frequent. Previous studies have shown a wide range of results. In a study in Denmark, 6.25% of respondents had wrist pain, and 5% had hand pain when training with an esports coach,^10^ similar to our findings. However, in another study in China, wrist pain was only reported in 8% of the study participants, whereas hand pain was reported in 38% of respondents who were professional mobile esports players.^11^ DiFrancisco-Donoghue et al. reported that 36% and 32% of collegiate esports players in the United States and Canada, respectively, experienced wrist and hand pain [9]. Fathuldeen et al. showed that 44.8% of competitive video gaming players in Saudi Arabia reported hand/wrist injuries.^12^ Unlike other body parts, the wrists and fingers require repetitive movements during esports activities. These repetitive movements may underpin wrist and finger pain. However, our study and these previous studies did not investigate repetitive movements of the wrist and finger, and thus, future studies should consider them.

The proportion of those with neck, wrist, and lower back pain and eye fatigue was significantly higher among professionals than amateurs. In traditional sports, professionals with periodized training schedules and access to team resources are less prone to musculoskeletal problems than amateurs.^15^ Our opposite result in esports players suggests an inadequate support system for professionals. Furthermore, regarding esports activities, professionals spent more time practicing on weekdays, were more likely to mainly use mobile devices, and were closer to the screen than amateurs. These three factors of esports activities were correlated with each other. Among them, the main device used and the distance between the screen and the face during esports activities were significantly associated with some physical complaints in the logistic regression analysis. Thus, these factors may have affected the high proportion of those with physical complaints among professionals.

Mainly using mobile devices and being closer to the screen and face during esports activities were associated with a higher proportion of those with neck pain, stiff or sore shoulders, lower back pain, and eye fatigue in the age-adjusted logistic regression analysis (Model 1). Although the importance of these studies has been noted, the proportion of those with physical complaints regarding the different devices used to play esports has not been examined.^20^ Some systematic reviews suggested associations between the regular use of mobile devices and physical complaints in the eyes, neck, and upper extremities.^23–25^ Furthermore, these two would be linked problems because there was a relatively high correlation between mainly using mobile devices and being closer to the screen during esports activities.

One of the reasons that these two esports activities factors were associated with physical complaints could be due to the player’s posture when playing esports. Mobile esports players are more likely to adopt a forward or flexed head posture for better eye focus, while the elbows press on the table to hold the arm and the mobile device. They may also have a slouched posture and rounded shoulders. Such poor posture can impose problems on the musculoskeletal and ocular systems.^26,27^

Based on our results, ergonomic approaches can effectively prevent physical complaints. Height-adjustable desks or desk-mounted armrests can help position the arm or mobile device higher or closer to the upper extremities. Increasing the strength and flexibility of the trunk muscles may also help address physical complaints. A previous study evaluated spinal function using an external noninvasive device and reported that professional mobile esports players demonstrated significantly poorer spine posture, mobility, and stability than the standard reference.^28^ Moreover, offering opportunities to stand during esports activities may efficiently alleviate physical complaints. While standing up in the middle of a set of games is difficult, providing opportunities to stand between sets is possible. Periodic interruption of sedentary behavior improves musculoskeletal symptoms.^29^ Taking a break while using electronic equipment also reduces computer vision syndromes, including eye fatigue.^30^

In a logistic regression analysis adjusted for age and esports level (Model 2), significant associations were only seen between the device mainly used and stiff or sore shoulders and between the distance between the screen and the face during esports activities and neck pain and stiff or sore shoulders. However, other odds ratios that showed significant associations in Model 1 remained high in Model 2. Thus, these associations may be strong. However, further studies are needed to target more esports players due to the small number of respondents in this study.

The duration of esports titles played primarily and hours of esports activities per day on weekdays and holidays had no significant association with physical complaints. A previous study reported that career duration was not significantly associated with ease of eyestrain or leg numbness,^11^ consistent with the findings of this study. However, another study in Saudi Arabia showed that the duration of playing video games was inversely related to musculoskeletal disorders, with gamers who played for 1–5 years showing a higher rate of having musculoskeletal disorders than those who played for more than 10 years.^12^ Also, Lindberg et al. reported that esports players with musculoskeletal pain participated in less esports training than those without pain.^10^ Indeed, excessive esports playing may negatively impact physical health. However, considering our study and previous studies, the findings of the cross-sectional studies cannot rule out the possibility that many esports players who are excessively playing are retired. Therefore, a longitudinal study is necessary to verify the causal relationships among these variables.

This study had some limitations. First, as noted above, since the number of respondents in this study was small, further studies are needed to target more esports players. Second, this study used cross-sectional data; therefore, we could not evaluate the causal relationship between esports activities and physical complaints. A longitudinal study is necessary to verify the causal relationships among these variables. Third, other factors related to esports activities should also be considered. For example, equipment characteristics and repetitive movements of the wrists and fingers may be related to physical complaints. Fourth, our study only included male players. Currently, there are a few female esports players.^31^ However, the number of women may increase in the future because physical attributes are unrelated to high performance in esports, allowing men and women to compete in the same events, unlike traditional sports.^32^ Therefore, it is necessary to accumulate knowledge about female esports players. Finally, data were collected using self-reported questionnaires. Therefore, reporter bias cannot be ruled out.

Despite these limitations, to the best of our knowledge, this is the first study to examine factors of esports activities from multiple perspectives related to physical complaints among esports players. Since esports players require long hours of esports activities to enhance their competitive performance, it is notable that we were able to identify clues to practical countermeasures for their physical complaints from factors other than the duration of esports activities.

## Conclusions

Esports players in Japan reported that stiff or sore shoulders were the most common complaint, followed by eye fatigue, headache, neck pain, and lower back pain. Mainly using mobile devices and being closer to the screen and face during esports activities were associated with a higher proportion of those with neck pain, stiff or sore shoulders, low back pain, and eye fatigue.

## Data availability statement

The data supporting the findings of this study are available from the corresponding authors upon reasonable request.

## Data Availability

The datasets used and analyzed in this study are available from the corresponding authors upon reasonable request.

## Acknowledgments

The authors would like to express their sincere gratitude to all the participants for their invaluable contributions to this study. Additionally, we would like to extend our heartfelt appreciation to the Ibaraki Esports Industry Creation Project, facilitated by Ibaraki Prefecture, Japan, for their indispensable support in fostering connections with esports teams. We are especially grateful to Mr. Okada, Mr. Kawamoto H. Ayato, and Mrs. Sawada, as well as the representatives from each esports team, for their invaluable assistance in participant recruitment, which played a vital role in the successful completion of our research.

## Authors contribution statement

Takafumi Monma conceptualized the study design, acquired, analyzed, and interpreted the data, and wrote the manuscript. Takashi Matsui conceptualized the study design, acquired, analyzed, and interpreted the data, and reviewed the manuscript. Shoya Koyama, Hiromasa Ueno, Junichi Kagesawa, Chisato Oba, Kentaro Nakamura, Hideki Takagi, and Fumi Takeda reviewed the manuscript. All the authors have approved the final manuscript.

